# Associations between maternal iron supplementation in pregnancy and offspring growth and cardiometabolic risk outcomes in infancy and childhood

**DOI:** 10.1101/2022.01.19.22269248

**Authors:** Clive J. Petry, Laurentya Olga, Ieuan A. Hughes, Ken K. Ong

## Abstract

It was previously observed that maternal iron supplementation in pregnancy was associated with increased offspring size and adiposity at birth, possibly mediated through increased risk of gestational diabetes. In this study we explored associations of maternal iron supplementation in pregnancy with postnatal offspring growth in infancy and indices of cardiometabolic disease risk factors such as glucose tolerance, insulin sensitivity and blood pressure in mid-childhood (at ∼9.5 years of age) in the Cambridge Baby Growth Study. In infancy adiposity-promoting associations with maternal iron supplementation in pregnancy were evident at 3 months of age (e.g. mean difference in skinfold thickness: β=+0.15 mm, p=0.02, in n=341 whose mothers supplemented versus 222 that did not; waist circumference: β=+0.7 cm, p=0.04, in n=159 and 78, respectively) but differences lessened after this time (e.g. 3-12 month change in mean difference in skinfold thickness: β=-0.2 mm, p=0.03, in n=272 and 178, respectively). At ∼9.5 years of age associations with maternal iron supplementation in pregnancy were not evident for markers of growth, glucose tolerance, insulin sensitivity or secretion. However, children whose mothers supplemented with iron in pregnancy had lower mean arterial blood pressures (β=-1.0 mmHg, p=0.03, in n=119 and 78, respectively). These results suggest that most of the associations of maternal iron supplementation in pregnancy on growth and adiposity evident at birth disappear during infancy, but there may be some evidence of long-term nutritional programming evident later in childhood.

## Introduction

In a contemporary birth cohort from a high-income country, we recently reported that maternal iron supplementation in pregnancy was associated with increased offspring size and adiposity at birth (in terms of weight, head circumference and specific skinfold thicknesses) [1], suggesting increased fetal growth. The mechanism of this enhanced growth could be linked to the parallel increased risk of the mother developing gestational diabetes (GDM) in pregnancy, potentially explaining previously reported findings relating to maternal multiple micronutrient supplementation in pregnancy [2]. The observed enhanced fetal growth in response to maternal iron supplementation in pregnancy is consistent with findings from randomised controlled trials which also reported iron supplementation leading to increased offspring birth weight [3, 4]. However, two systematic reviews failed to find significant overall effects of maternal iron supplementation in pregnancy on offspring birth weight [5, 6].

Maternal iron status in pregnancy can affect the offspring post-partum. It has been speculated that *in utero* exposure to iron supplementation in pregnancy may also have long-term consequences for the offspring [7]. Investigations looking at long-term effects of altered maternal iron intakes in pregnancy in human offspring are scarce. However, studies in pregnant rats have shown long-term effects of restricted dietary iron including raised blood pressure [8, 9] and lower circulating glucose concentrations [8] in the offspring who were fed iron-replete diets following weaning [9-12]. The focus on iron deficient diets in rats reflects the fact that worldwide iron deficiency is the most prevalent form of malnutrition in humans. Effects of iron supplementation in rat pregnancies on the offspring are less commonly studied. However, maternal iron supplementation in murine pregnancy has been reported to increase body weight of the offspring at birth [13] and at weaning [14]. This is consistent with the lower birth weights in the maternal iron restriction rodent models [8, 9], and raises the possibility that maternal iron supplementation in pregnancy might also lead to long-term effects in the rat offspring on blood pressure and glycaemia.

Long-term effects of maternal iron deficiency on the rat offspring have been described as an example of nutritional programming [9], defined as the process through which variation in the quality or quantity of nutrients consumed during pregnancy exerts permanent effects upon the developing fetus [15]. Although evidence of nutritional programming has been extensively observed in animal models, in humans it has been more limited because many relevant studies were retrospective, and their results could not establish causality [16, 17]. However, a single prospective study of the long-term effects of maternal iron supplementation in pregnancy from a high-income country found an association with lower systolic blood pressure in children aged 10 years of age [18]. This association was attenuated when adjusting for cord serum ferritin concentrations. This implies that the reduction in blood pressure may have been due to nutritional programming related to the maternal iron supplementation, or at least iron levels, in pregnancy.

The present study was performed to evaluate associations between maternal iron supplementation in pregnancy and markers of offspring growth in infancy and cardiometabolic risk factors in mid-childhood. We did this using the prospective Cambridge Baby Growth Study (CBGS), which has extensive infancy growth records and has recently been extended by following a subset of the participants into mid-childhood [19].

## Methods

### Cambridge baby growth study

A total of 2,229 pregnant women were recruited to the CBGS from (∼week 12) ultrasound clinics held at the Rosie Maternity Hospital, Cambridge, U.K. between the years 2001 and 2009. At recruitment all the women were given a pregnancy questionnaire [1, 2] to fill in as the pregnancy progressed, with help offered from research nurses if required. These questionnaires were collected post-partum. After 571 of the pregnant recruits withdrew from the study prior to the birth of their babies, this left 1,658 women to participate in this initial phase of the study, along with their babies and partners who were subsequently also recruited to the study. Over 95% of the babies recruited were classified as being White, the remainder being Black (African or Caribbean) (1.3%), Asian (1.7%) or mixed race (1.7%). Ethical approvals for the CBGS and its follow-on study were granted by the Cambridge (Local) Research Ethics Committee: LREC Ref: 00/325, REC Ref: 08/H0302/47. The child’s legal guardian provided written informed consent for both phases of the study, whereas the child themselves only provided it for the follow-up phase.

*A priori e*xclusion criteria for this analysis were: study withdrawal prior to delivery, missing pregnancy questionnaires, especially the question about dietary supplementation in pregnancy [1], and this pregnancy leading to the birth of more than one baby (because of the impact of multifetal pregnancies on offspring growth). The maximal sampling frame for this analysis was 898 participants who had anthropometric and skinfold measurements at 3 months of age (with 291 mothers who completed an additional temperament questionnaire at this age), 802 participants at 12 months and 730 at 24 months, plus 201 participants in the follow-up study of mid-childhood. Associations with measurements of the offspring at birth have already been reported [1].

### Assessment of maternal iron supplementation in pregnancy

For this analysis women were classified as either having supplemented their diets with iron in pregnancy or not in the same manner as that used previously [1], i.e. via the response to the question in the pregnancy questionnaire about whether or not the mother took supplements in pregnancy. Data was also collected about diseases that the participants may have suffered from in pregnancy; responses indicating any form of anaemia were coded as ‘anaemia’ for this analysis. Although 1,239 women returned completed pregnancy questionnaires after the birth of their babies, not all of these were included in this analysis due to factors relating to exclusion criteria or not providing clear answers to the question about pregnancy supplements.

### Infancy growth assessments

In the present analysis birth assessments in the offspring were used to calculate changes in growth between assessments (i.e. 0-3, 3-12 and 12-24 months of age). Offspring birth weights were documented from hospital records. Other “newborn” measurements, that included length, head circumference and skinfold thicknesses, were made in triplicate by one of three trained paediatric research nurses within the first 8 days of life. The same set of measurements were also made on the infants at around 3, 12 and 24 months of age by the same nurses. From September 2006 the infants’ waist circumferences were also measured in the children who were 3 months of age or older [21]. Weight was measured using a 757 Baby Scale, and length using a 416 Infantometer (both from SECA, Birmingham, U.K.). Head and waist circumferences were evaluated using a standard tape measure. Skinfold thickness was measured in triplicate at four sites (flank, quadriceps, subscapular, triceps) on the left side of the body using a Holtain Tanner/Whitehouse Skinfold Calliper (Holtain Ltd., Crymych, U.K.) as described [20].

### Follow-up assessments

Between September 2013 and October 2018, 817 invitation letters to attend a research clinic were sent out to children who had been assessed in infancy and who were by then 5-11 years of age [19]. All these children were born at a gestational age of greater than 33 weeks and had been assessed during infancy at 12 months of age and/or beyond. Those that were excluded from being invited (having checked the National Health Service Information Centre for health and social care resource) included those that had another serious illness or had died, those that used a high dose glucocorticoid medication (oral or inhaled), those that had a congenital or genetic growth disorder, and those that had previously withdrawn from the study. If there was no response to the invitation after a month a reminder letter was sent. Of all the children that were invited 365 responded, and 285 consented and took part in the follow-up study. Of these 122 had mothers that supplemented their diets with iron in pregnancy and 79 had mothers that did not.

Participants in the follow-up study fasted overnight (> 8 hours) before attending the hospital’s clinical research facility the following morning. Weight was measured using electronic scales with the child wearing light clothing without shoes. Height was determined using a wall-mounted stadiometer. Skinfold thicknesses were measured in the same manner that they had been in infancy. Blood pressure and resting pulse rate were quantified as the mean of 3 readings using an automated sphygmomanometer. Glucose tolerance and insulin sensitivity and secretion was assessed using a short (30 min.) 1.75 g/kg body weight (maximum 75 g) oral glucose tolerance test (OGTT). Blood samples (capillary and venous for the measurement of glucose, insulin and C-peptide concentrations) were collected in the fasting state and then again, 30 mins. after the oral consumption of the glucose load.

### Laboratory assays

Plasma insulin and C-peptide concentrations in mid-childhood were measured using in-house Meso Scale Discovery immunoassays, with both intra- and inter-assay variation <5 % throughout. Blood glucose concentrations were measured on venous samples using a routine glucose-oxidase method (Yellow Springs Instruments, Yellow Springs, Ohio, USA).

### Calculations

Changes in the various growth assessments were calculated by subtracting the measurements at the younger age from those made at the older age. The body mass index (BMI) was calculated as the reciprocal of the height squared multiplied by the weight. Insulin resistance was assessed using the fasting C-peptide concentrations and HOMA IR. HOMA IR was calculated from fasting C-peptide and glucose concentrations using the HOMA Calculator [21]. Insulin secretion was assessed using the insulin increment, the insulinogenic index and the insulin disposition index. The insulin increment was calculated as the OGTT 30 min. insulin concentration minus the fasting concentration. The insulinogenic index was calculated as the insulin increment divided by the rise in glucose concentrations over the 30 mins. of the OGTT. The insulin disposition index was calculated as the insulinogenic index divided by the C-peptide derived HOMA IR. The mean arterial blood pressure was calculated as a third of the sum of twice the diastolic blood pressure plus the systolic blood pressure [22]. The pulse pressure was calculated by subtracting the diastolic blood pressure from the systolic blood pressure. The index of multiple deprivation (IMD) was estimated from the participant’s residential postcode [23]. The mean skinfold thickness of each individual was calculated as a quarter of the sum of the measurements at four skinfold sites. Internal z-scores were calculated by dividing the residuals from a regression model, including age and sex, by the cohort standard deviation.

### Statistical analyses

Associations of maternal iron supplementation in pregnancy with offspring growth and development at single timepoints (ages 3, 12 and 24 months) were tested by multiple linear regression. The models included the potential confounders: sex, age at assessment, gestational age at birth (only included at 3 months of age), maternal ethnicity, maternal pre-pregnancy BMI, maternal height (in models for offspring length or height), maternal smoking in pregnancy, parity, socioeconomic status (as reflected by the IMD) and breastfeeding history at 3 months of age. These confounders were chosen *a priori* as correlates of offspring growth in the CBGS [20] and ALSPAC [24]. A further analysis was performed on subscapular skinfold thickness measurements, using multiple linear regression to compare results from those offspring whose mothers took iron supplementation in pregnancy that was not part of multiple micronutrient preparations with those with mothers who did not supplement their diets with iron in pregnancy. Subscapular skinfold thickness measurements were used as they were the one measurement that had maintained a significant difference at birth (despite the reduction in statistical power) when those mothers who supplemented their diets with multiple micronutrient preparations were excluded from previous analyses [1].

The follow-up study was also analysed using multiple linear regression. The covariates in the models were age and sex, with the arm used to measure blood pressure in added where appropriate. A sensitivity analysis was performed by assessing associations between maternal iron supplementation status in pregnancy and results from the follow-up study just in those children whose mothers did not report anaemia in pregnancy in their questionnaire.

Statistical analyses were conducted using Stata (version 13.1; StataCorp LP, College Station, Texas, U.S.A.) and R (version 3.3.2; R Foundation for Statistical Computing, Vienna, Austria). Data are mean (95% confidence interval) unless stated otherwise. p < 0.05 was considered statistically significant throughout.

## Results

### Clinical characteristics of study participants

The characteristics of the participants that were included in the infancy analysis were similar to those of the CBGS participants that were not included (Table 1). However, the gestational age at birth was longer in the participants by around two days (as the non-participants included offspring of twin pregnancies, who generally were born with lower gestational ages at birth) and fewer of their mothers reported smoking during pregnancy. The characteristics of the follow-up study participants were largely representative of the bigger group of people that took part in the infancy analysis, except for the fact that there was a different proportion of boys to girls (Table S1).

**Table 1.** Characteristics of those participants (of the infancy growth studies) in this analysis and those participants from the Cambridge Baby Growth Study who did not participate in this analysis.

| Clinical characteristic | Participants | Non-participants | p-value |
| --- | --- | --- | --- |
| Mother supplemented their diet with iron in pregnancy (n yes/no) | 593/392 | 13/3 | 0.1 |
| Gestational age at birth (weeks) | 39.9<br>(39.8, 40.0)<br>(n=968) | 39.6<br>(39.5, 39.7)<br>(n=689) | 9.3x10<sup>-4</sup> |
| Birth weight (kg) <sup>1</sup> | 3.491<br>(3.462, 3.519)<br>(n=964) | 3.454<br>(3.420, 3.488)<br>(n=687) | 0.1 |
| Birth length (cm) <sup>2</sup> | 51.4<br>(51.3, 51.6)<br>(n=938) | 51.3<br>(51.2, 51.5)<br>(n=656) | 0.2 |
| Head circumference at birth (cm) <sup>2</sup> | 35.3<br>(35.2, 35.4)<br>(n=940) | 35.3<br>(35.2, 35.4)<br>(n=657) | 0.4 |
| Ponderal index at birth (kg/m<sup>3</sup>) <sup>2</sup> | 25.6<br>(25.5, 25.8)<br>(n=936) | 25.5<br>(25.3, 25.7)<br>(n=655) | 0.5 |
| Sex (males/females) | 505/461 | 648/630 | 0.5 |
| Mother smoked in pregnancy (n<br>yes/no) | 31/936 | 55/633 | 1.5x10<sup>-5</sup> |

Data are numbers of CBGS participants or means (95% confidence intervals).
<sup>1</sup>adjusted for gestational age at birth and sex.
<sup>2</sup>adjusted for gestational age at birth, age at assessment and sex.

### Growth and development in infancy

At 3 months of age infants whose mothers reported supplementing their diets with iron in pregnancy had increased (mean ± SD) skinfolds thickness z-scores (maternal iron supplementation 0.08 ± 0.80 (n=341), no maternal iron supplementation -0.07 ± 0.76 (n=222), adjusted p=0.02) and waist circumferences (maternal iron supplementation 41.3 ± 2.8 cm (n=159), no maternal iron supplementation 40.6 ± 2.5 cm (n=78), adjusted p=0.04) in comparison to those whose mothers did not supplement. There was no difference in weight, height, BMI or head circumference (data not shown). At 12 months of age only the waist circumference showed a significant difference (maternal iron supplementation 46.0 ± 3.1 cm (n=174), no maternal iron supplementation 45.0 ± 3.2 cm (n=109), adjusted p=0.003). By 24 months of age none of the anthropometric measurements differed according to maternal iron supplementation group. In terms of the changes between the different timepoints, the significant difference was a reduction in mean skinfold thickness z-scores between 3 and 12 months of age in those children whose mothers reported supplementing with iron in pregnancy (maternal iron supplementation -0.08 ± 0.92 (n=272), no maternal iron supplementation 0.12 ± 0.88 (n=178), adjusted p=0.03) (Fig 1).

**Fig 1.**
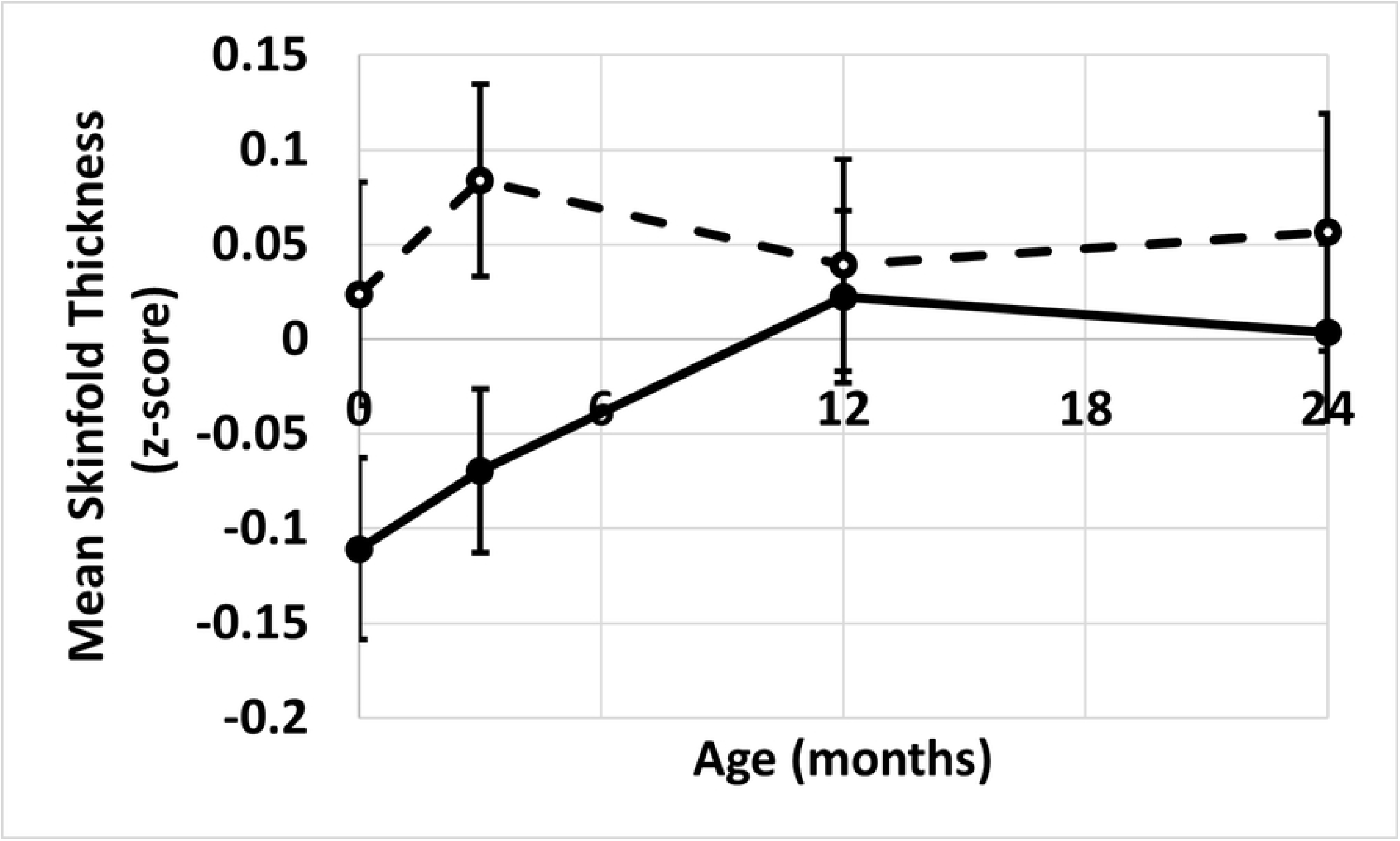
The change in mean skinfold thickness z-score over the first two years of life.

The dotted line represents data from children (n=272) whose mothers supplemented with iron during pregnancy. The solid line represents data from children (n=178) whose mothers did not report supplementing with iron during pregnancy. Data are mean (SEM).

At 3 months of age the subscapular skinfold thicknesses were larger in offspring of women who supplemented their diets with iron in isolation (maternal iron supplementation 7.4 ± 0.7 mm (n=12), no maternal iron supplementation 6.9 ± 0.6 mm (n=44), adjusted p=0.048). This borderline significant difference was still evident at 12 months of age (maternal iron supplementation 7.6 ± 0.8 mm (n=11), no maternal iron supplementation 7.1 ± 0.6 mm (n=37), adjusted p=0.04) but had disappeared by 24 months of age (maternal iron supplementation 6.6 ± 0.6 mm (n=9), no maternal iron supplementation 6.5 ± 0.5 mm (n=32), adjusted p=0.5).

### Mid-childhood follow-up

There was no detectable difference according to maternal iron supplementation in pregnancy in any of the anthropometric measurements made in mid-childhood (Table 2). There were also no detectable differences in the circulating glucose concentrations or in the markers of insulin secretion or sensitivity. There were differences in blood pressure, however. Children whose mothers supplemented with iron in pregnancy had lower mean (95% confidence interval) arterial blood pressures (β=-1.0 (−1.0, -0.9) mmHg, adjusted p=0.03, in n=119 whose mothers supplemented and 78 that did not). Similar results were evident for diastolic blood pressure (β=-2.1 (−4.1, -0.1) mmHg, adjusted p=0.04). A non-significant association in the same direction was found with systolic blood pressure (β=-2.8 (−5.9, 0.3) mmHg, adjusted p=0.07).

**Table 2.**
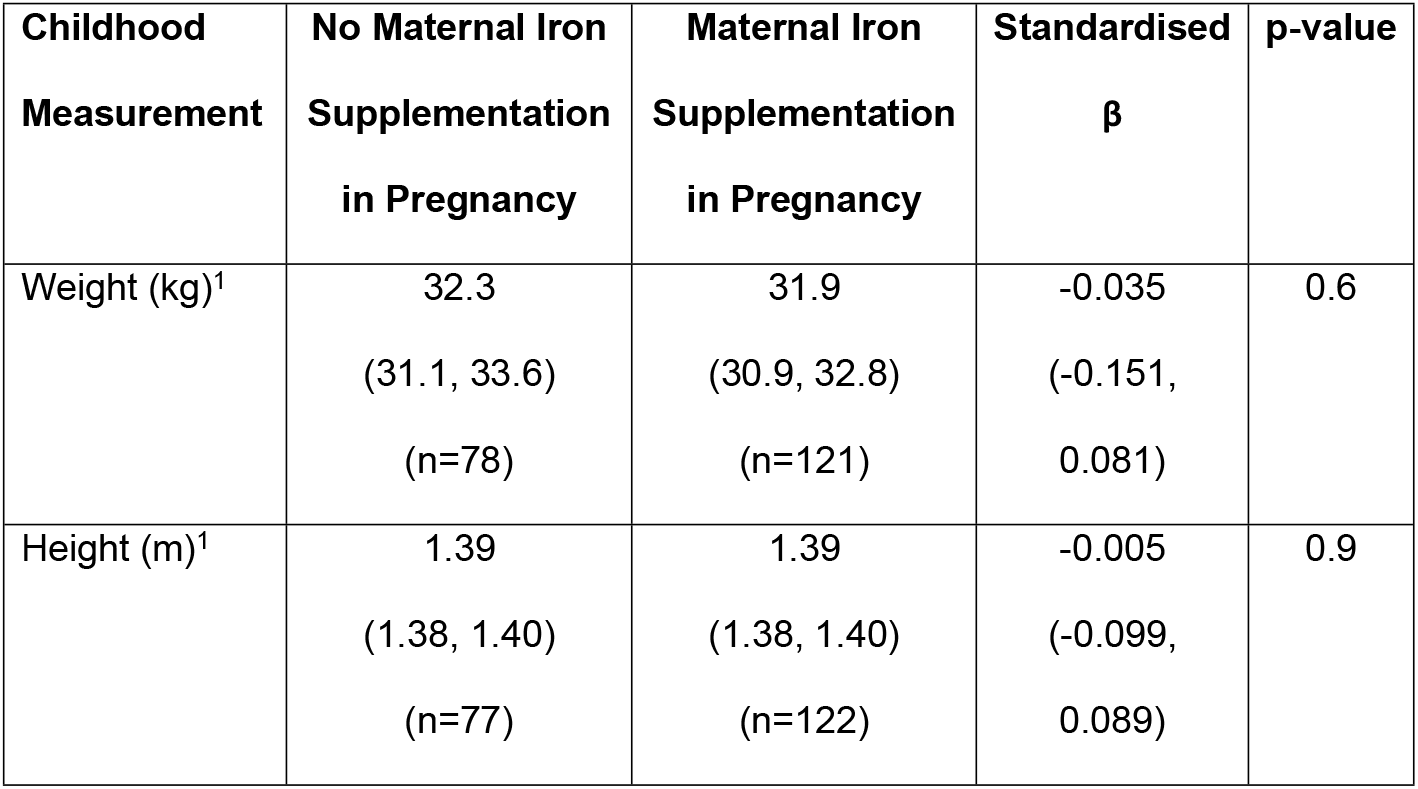
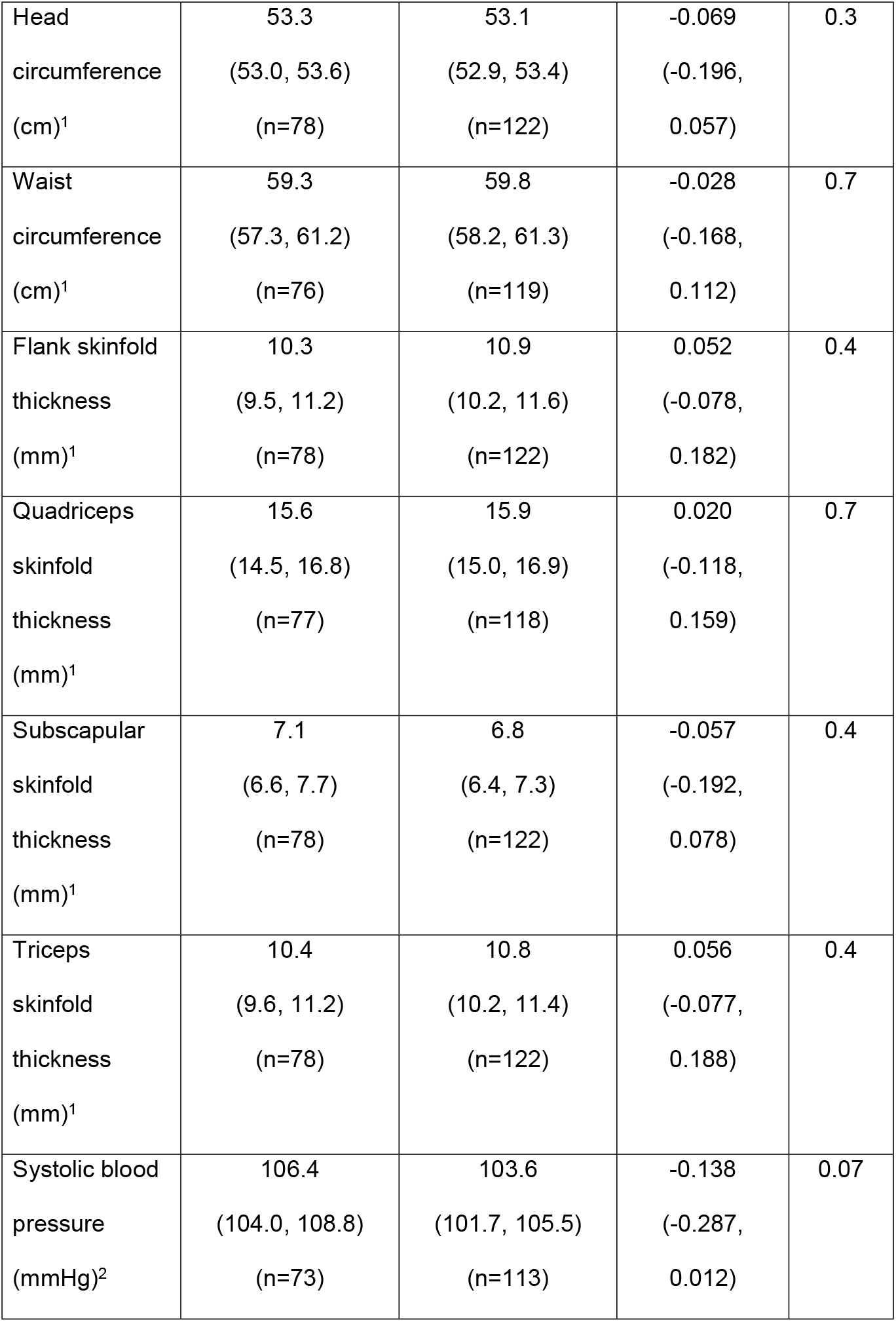
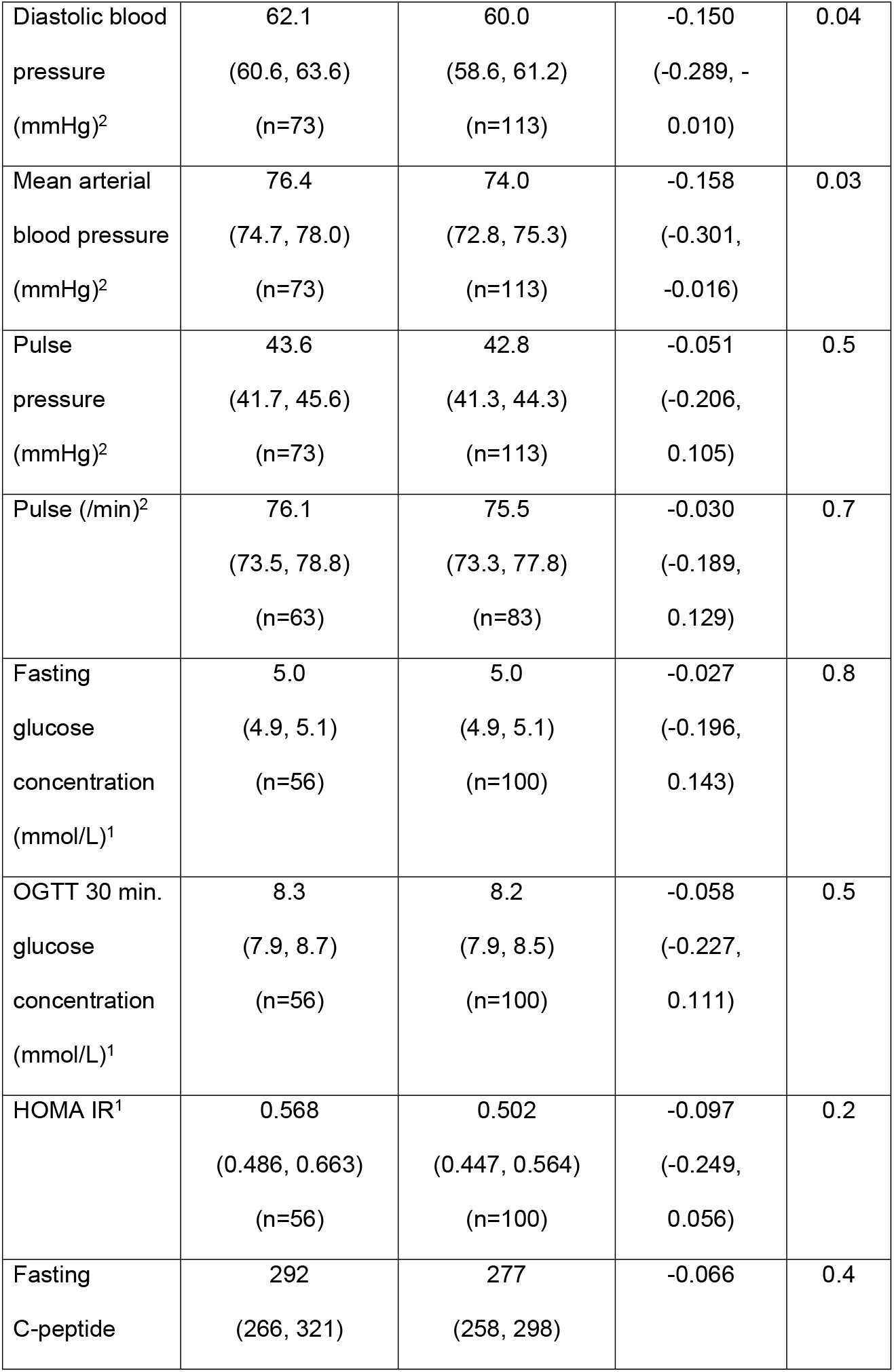
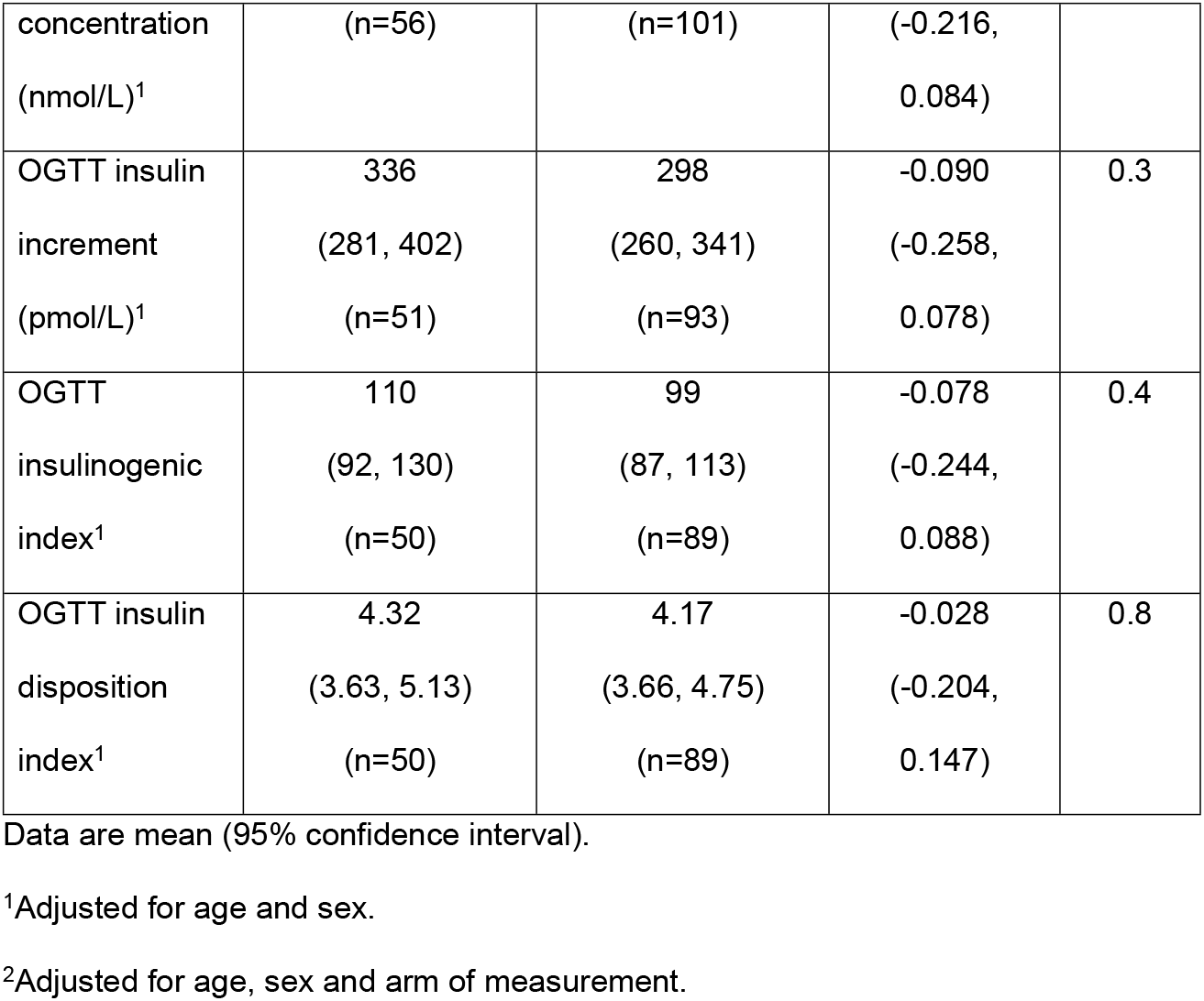
Associations between maternal iron supplementation status in pregnancy and offspring growth and physiological measurements at around age 9.5 years.

A sensitivity analysis, excluding children whose mothers reported anaemia in pregnancy, produced similar results (Table S2). Thus children (aged ∼9.5 years) whose mothers supplemented with iron in pregnancy had lower mean arterial blood pressures (β=-1.0 (−1.0, -0.9) mmHg, adjusted p=0.04, in n=103 whose mothers supplemented and 70 that did not) and diastolic blood pressures (β=-2.1 (−4.2, -0.1) mmHg, adjusted p=0.04).

## Discussion

The results of this analysis indicate that the previously reported increases in size and adiposity at birth of babies whose mothers had supplemented their diets in pregnancy with iron [1] gradually disappear in infancy. In mid-childhood most of the physiological measurements were also not associated with maternal iron supplementation in pregnancy with the exception of (mean and diastolic) arterial blood pressures, which were lower in those children whose mothers supplemented with iron in pregnancy. These data are consistent with the idea that maternal iron supplementation in pregnancy may programme blood pressure in the offspring but not overall size and adiposity.

Previously we reported that maternal iron supplementation in pregnancy was associated with increased weight, head circumference, BMI and skinfold thicknesses in the offspring at birth [1]. In the current analysis these increases attenuated with time, albeit with different measures doing this at different rates. The first potential explanation for the reduction in effect sizes associated with maternal iron supplementation in pregnancy is that the increases in size and adiposity at birth relate to the increased risk of maternal GDM [1], and that, with the cessation of this drive postnatally, associated effects are just residual. The rates of attenuation of associations with separate measures then differ as a number of other factors are involved in overall and regional body growth. The second potential explanation for the reduction in effect sizes associated with maternal iron supplementation in pregnancy relates more directly to infant iron stores. It is believed that iron needs for the first six months of life are met by stores present at birth [25]. It is therefore not necessarily surprising that in the present analysis at 3 months of age there was still evidence of increased adiposity in terms of skinfold thicknesses and waist circumference in those children whose mothers supplemented their diets with iron in pregnancy, even if the increases in weight and head circumferences had attenuated. By around 6 months of age, with the stores from birth now being depleted, iron needs to be topped up from solid food and/or iron-enriched formula (breast milk containing insufficient iron [26]), so effects of maternal iron supplementation in pregnancy that did not involve programmed changes in cellular structure or physiological processes would be expected to have diminished. Consistent with this concept, in our analysis between 3 and 12 months of age the effect of maternal dietary iron supplementation in pregnancy on skinfold thicknesses attenuated, such that at 1 year of age only the waist circumference was associated with maternal iron supplementation in pregnancy. No other effects were evident, as has been observed from 6 months of age in two previous studies [27, 28]. By 2 years of age in the present study even the association with waist circumference observed at 1 year of age was no longer evident. Whichever of the potential explanations is the correct one, the growth patterns observed in the current analysis suggest that the increased size and adiposity at birth associated with maternal iron supplementation in pregnancy [1] are not due to associated long-term programming effects, but instead may result somehow from associated short-term alterations like maternal-influenced iron stores or the development of maternal GDM [29].

Not surprisingly, given the results from infancy, the anthropometric measurements in mid-childhood failed to show an association with maternal iron supplementation in pregnancy. Similarly, most of the physiological measurements did not show an association. However, consistent with results from 10 year olds in ALSPAC [18], blood pressure was lower in children aged around 9.5 years of age whose mothers supplemented with iron in pregnancy. This lowering of blood pressure was evident in the mean arterial and diastolic blood pressures, with systolic blood pressure altering in the same direction, albeit non-significantly. In a randomised clinical trial conducted in low birth weight children, iron supplementation in infancy was also associated with reduced blood pressure at age 7 [30]. These results could also be relevant to results from the present analysis given that iron stores established in fetal life are relied upon pre-weaning to prevent iron deficiency in the first 6 months of life [25].

Long-term effects of maternal iron manipulation in pregnancy on offspring blood pressure but not glucose tolerance are also somewhat consistent with results from the rat models [9], even if they were in the opposite direction related to the fact that the rats’ mothers were fed an iron deficient diet in pregnancy rather than their diets being supplemented with it. The mechanism of the nutritional programming leading to blood pressure changes in the rat models appear to be at least partially related to altered intrarenal haemodynamic properties [11] and/or changes in numbers of nephrons [31]. It is possible that alterations in these renal factors underpin the subtle changes in blood pressure that we observed in humans, although the literature is devoid of evidence to that effect, and maternal iron supplementation was not associated with kidney volume in their children in one study [32]. Intriguingly though, in the MINIMat trial in rural Bangladesh, the glomerular filtration rate in children aged around 4.5 years was higher in those whose mothers had received 60 mg iron supplements in pregnancy compared with those whose mothers received half that dose [33]. Somewhat inconsistent with both our observation and that in ALSPAC [18], however, there was no association between level of maternal iron supplementation in pregnancy and blood pressure in childhood in this study.

This analysis has a number of limitations. Firstly, we do not have access to the iron status of the study participants, either the mothers during pregnancy or their children. Thus, effects associated with maternal iron supplementation could have been confounded by dietary iron deficiency in pregnancy or effects established by other components of multiple micronutrient populations. However, results from our sensitivity analyses, where we found similar associations in mid-childhood in offspring of women who did not report having anaemia during pregnancy to those of the full analysis, are inconsistent with the former possibility. In contrast to our findings at birth [1], increased iron status in infancy appears to be negatively related to growth [34, 35]. As maternal and infant iron statuses may be related [36] a high iron status (resulting from maternal iron supplementation in pregnancy) may explain both an increased size and adiposity at birth and a convergence to growth trajectories of those whose mothers did not supplement with iron in pregnancy, in infancy. Further studies are needed to clarify this, however. Although other non-iron components of multiple micronutrient preparations could have confounded our analyses, at the birth of the offspring we found that significant associations with maternal multiple micronutrient supplementation were completely attenuated by adjusting for maternal iron supplementation [1] and associations with iron taken in isolation were all in the same direction as those observed with multiple micronutrient supplementation (albeit not significantly in the reduced number of participants). Also, in the present analysis associations between iron supplementation specifically (with exclusion of those whose mothers supplemented with multiple micronutrient preparations) and offspring subscapular skinfold thickness showed a similar growth pattern to those observed when including those infants whose mother’s iron supplementation was as part of multiple micronutrient preparations. Also, the significant association with lower offspring blood pressures in the present analysis is consistent with the findings in ALSPAC [18], where the significant association was attenuated when adjusted for maternal circulating ferritin concentrations. These factors therefore suggest that iron supplementation specifically could have mediated the significant associations in the present study. A second limitation of this study is that the follow-up study participants may not be fully representative of the cohort as a whole, especially with regard to the differences in proportions of boys to girls. This could have biased the results. However, the most important results from mid-childhood in this analysis relate to blood pressure and as there were no sex-related differences in blood pressure in our cohort (data not shown) our main conclusions are unlikely to have been affected by any such biases. A third limitation of the study is its potential limited relevance to other populations given that the cohort was recruited from a single centre and is almost exclusively White Caucasian [20]. Further prospective studies, particularly those using cohorts of other ethnicities, are required to corroborate the relevance of our findings to other populations.

In conclusion, this analysis suggests that increased growth and adiposity at birth resulting from maternal iron supplementation in pregnancy does not result from nutritional programming *per se*, at least in a high-income setting, as in infancy there is a convergence to the size and adiposity of those whose mothers did not supplement with iron in pregnancy. However, blood pressure in mid-childhood may be programmed since the current analysis and the one previous study that investigated this [18], found lower blood pressure in children whose mothers had supplemented their diets with iron in pregnancy ten years previously. It is not currently clear whether the critical time period of this programming is pregnancy or infancy, however, since iron used during the first 6 months of life are largely stored from pregnancy [28, 29] and iron supplementation during this time also leads to reduced blood pressure in mid-childhood [30].

## Data Availability

All data produced are available online at https://doi.org/10.17863/CAM.79743

## Acknowledgments

The authors gratefully acknowledge the absolute importance of the contributions made to this study by Dr. Carlo Acerini and Prof. David Dunger, who were Principal Investigators of the Cambridge Baby Growth Study prior to their deaths in May 2019 and July 2021, respectively. We also acknowledge the excellent technical assistance for this project that was provided by Dianne Wingate and Karen Whitehead (University of Cambridge Department of Paediatrics). The authors would like to thank all the families that took part in the Cambridge Baby Growth Study, and acknowledge the crucial role played by the research nurses especially Suzanne Smith, Ann-Marie Wardell, and Karen Forbes (all University of Cambridge, Department of Paediatrics), staff at the Addenbrooke’s Wellcome Trust Clinical Research Facility, and midwives at the Rosie Maternity Hospital.

## Author contributions

CJP and KKO were involved in the project conception. IAH and KKO gained funding for the CBGS. CJP and LO performed the data analysis. CJP, LO, IAH and KKO were involved in the writing and revising of the manuscript.

## Availability of data

All data used for this analysis are available at https://doi.org/10.17863/CAM.79743.

## Supporting information

**<u>Table S1</u> Characteristics of those participants who took part in both the infancy and follow-up study and those participants that only took part in the infancy studies**.

**<u>Table S2</u> Associations between maternal iron supplementation status in pregnancy and offspring growth and physiological measurements at around age 9.5 years in those children whose mothers did not report being anaemic in pregnancy**.

